# Age-Dependent Sex Disparities in Sjogren’s Disease Prevalence Align with Natural Hormone Fluctuations

**DOI:** 10.1101/2025.02.26.25322717

**Authors:** Eliza C. Diggins, Melodie L. Weller

## Abstract

Sjogren’s Disease (SjD), like many autoimmune diseases, shows a strong female-to-male sex bias, raising the possibility of a hormone-mediated mechanism. However, specific hormonal drivers and the underlying mechanisms remain unclear. In this study, we analyze a electronic health record dataset containing >100,000 SjD patients, allowing for a comprehensive statistical investigation into sex bias and serum hormone levels associated with SjD susceptibility. Focusing on testosterone, estradiol, and sex hormone-binding globulin (SHBG), our analysis reveals an age-dependent trend in SjD’s sex bias, with a substantially reduced female bias observed in pediatric cases. This finding, alongside statistical modeling, suggests that testosterone or estradiol may influence the sex disparity in SjD prevalence, with distinct effects at different life stages. Specifically, we observe a higher percentage of male SjD patients in early childhood (30.06%, [CI: 26.16-34.11]), declining sharply in late pubescence into adulthood (9.85% [CI: 9.51–10.16]), followed by a secondary steady increase in male prevalence among older adults (13.50% [CI: 13.25-13.76]). Our models indicate that these shifts in sex bias align with typical age-related hormonal changes rather than abnormal levels, suggesting that natural hormone fluctuations may play a significant role in modulating disease susceptibility. This research not only refines our understanding of SjD but also underscores the importance of hormone-level monitoring in future SjD studies and potential therapeutic strategies. Advanced studies are required to further clarify whether testosterone or estradiol is the primary mediator of these trends, promising new insights into hormone-linked pathways and autoimmune disease pathogenesis.

## Introduction

Sjogren’s disease (SjD) is a complex autoimmune disorder characterized by immune-mediated damage to mucosal tissue and moisture-secreting glands, resulting in dry eyes and mouth, reduced tear production, and various glandular and extraglandular symptoms^1–4^. SjD also exhibits a strong female sex bias, with ≥85% of patients being female, raising the hypothesis of a hormone-mediated mechanism contributing to disease susceptibility^5,6^. However, the mechanisms underlying this sex bias remain unclear.

Previous studies have reported mixed findings regarding hormone abnormalities in SjD patients compared to controls, posing challenges in defining hormone-driven etiological factors^7–11^. Here, we propose a dynamic model where age-related hormonal fluctuations, rather than abnormal hormone levels, modulate sex-specific disease susceptibility over the lifespan.

Using >100,000 SjD electronic health records (EHR) and 1.4 million non-SjD controls from TriNetX Research Network, combined with NHANES baseline hormone data, we analyzed the relationship between sex-specific prevalence, hormone profiles, and age. Our findings reveal striking age-dependent shifts in SjD sex bias, particularly in pediatric and older populations, emphasizing the need for further research into hormone-associated immune modulation in SjD pathogenesis.

## Methods

We analyzed sex-specific prevalence and hormone associations in SjD using patient records from the TriNetX Research Network (TNXRN) and baseline hormone data from NHANES. Details on cohort selection, data processing, and statistical methodologies, including male sex prevalence calculation and hormone imputation, are provided in the **Supplemental Methods**. The study was conducted with ethical approval from the Institutional Review Board (IRB) at the University of Utah.

## Results

We analyzed 101,856 Sjogren’s disease (SjD) patients and over 1.3 million non-SjD controls identified from the TNXRN dataset to investigate sex-specific and age-dependent trends in disease prevalence (**Table 1**). Of the 101,856 SjD patients, 540 pediatric patients (ages 0–15) were identified, with 33 having available hormone data. The next age group (15–52.5) included over 3700 individuals with hormone data, and hormone testing rates remained low but consistent across all age groups. Analysis of the age-dependence in male SjD prevalence indicated a dynamic interaction between hormone levels and disease susceptibility across the lifespan (**Figure 1**). Male prevalence peaks in pediatric patients (ages 5–15) at 30.06% [CI: 26.16–34.11%], declines to its lowest in adults (ages 15–52.5) at 9.84% [CI: 9.51–10.16%], and rises again in older patients (ages 52.5+) to 13.50% [CI: 13.25–13.76%] (**Figure 1A-B**). This trend reflects age-related modulation, with a decrease of -2.3% per year during pediatric ages and a gradual increase of 0.36% per year in older adults. The sex prevalence fraction (SPF) curve closely mirrors age-related fluctuations in serum testosterone and estradiol levels, emphasizing the influence of hormonal changes on disease susceptibility.

**Table 1.**
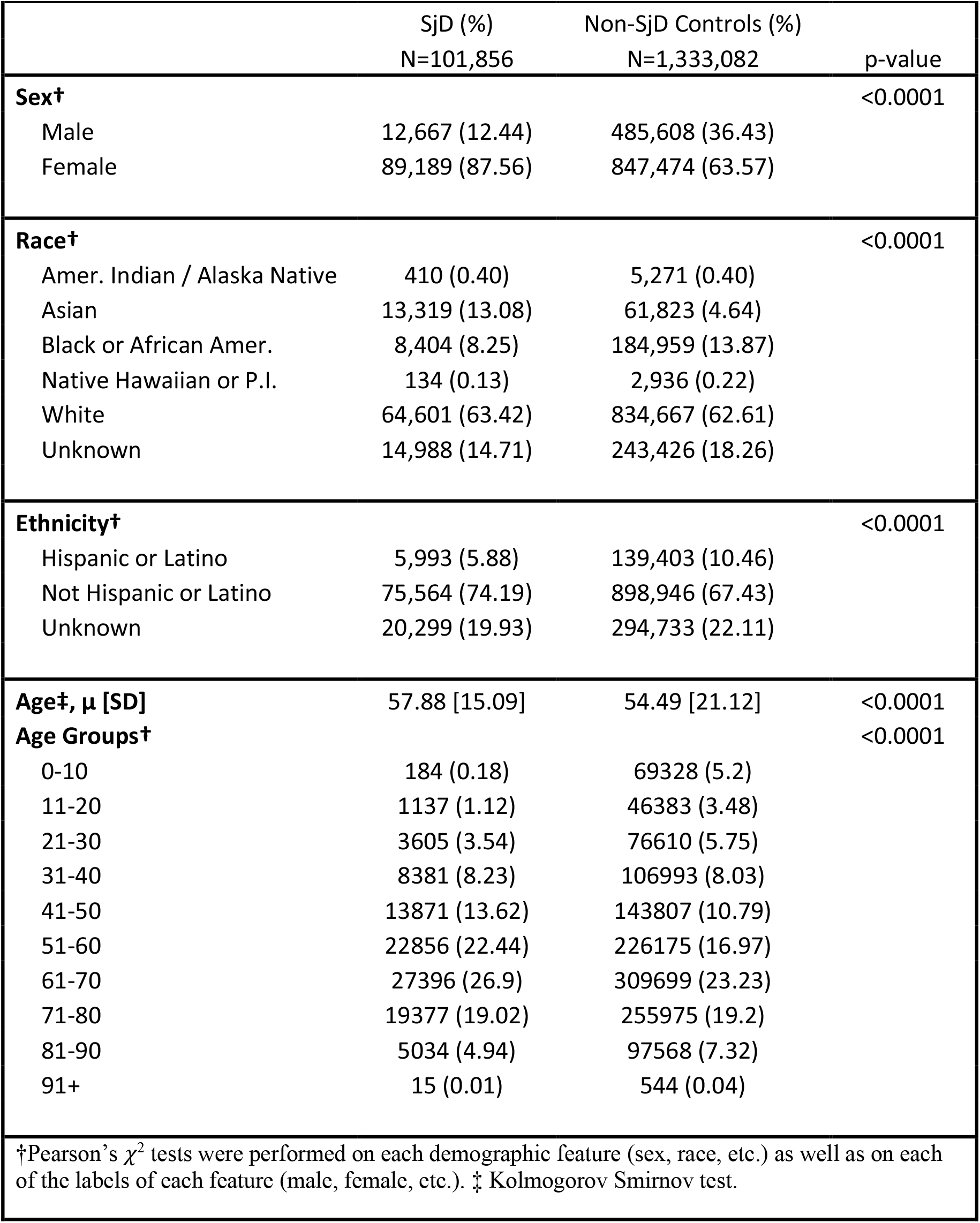
Demographic comparison of SjD and control cohort.

**Figure 1:**
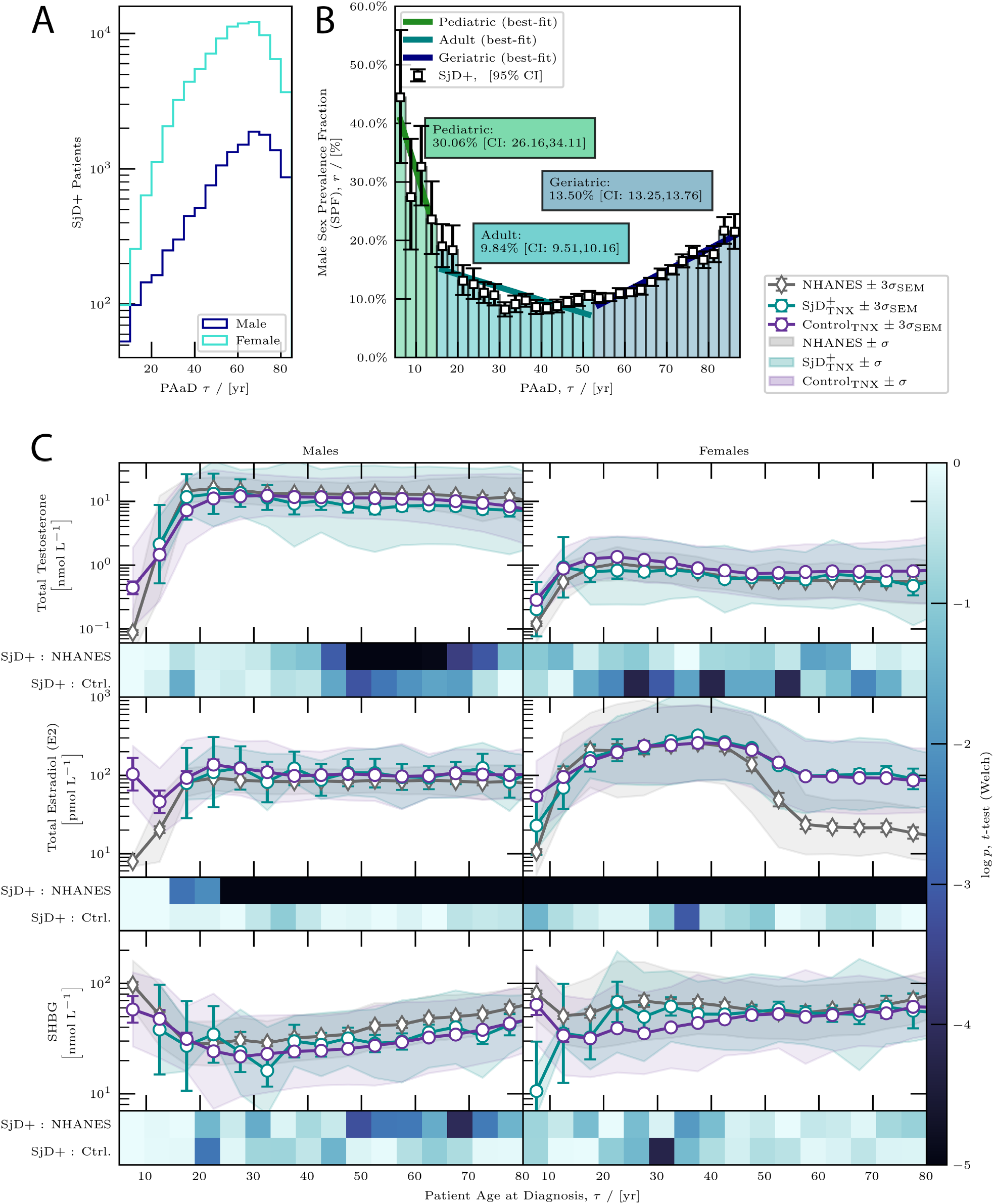
Age-Based Sex Bias and Serum Hormone Levels in SjD. **(A)** Age-based male sex prevalence (SPF) in SjD and **(B)** timeline of SjD diagnosis with 95% credible intervals, shown in 2.5-year age bins from ages 5 to 90. Categories include pediatric (<15 years), adult (15–52.5 years), and geriatric (>52.5 years). Best-fit OLS lines for each group indicate trends, with slopes of -2.29%, -0.22%, and 0.36% per year, respectively, calculated using Bayesian methods. **(C)** Mean serum hormone levels (SHLs) for testosterone, estradiol, and sex hormone binding globulin (SHBG) across SjD, control, and NHANES cohorts, displayed by sex. Heatmaps show ***p***-values (Welch’s ***t***-test) for differences across bins, with SHLs modeled as log-normal and MLE applied for means. Error bars represent 3***σ*** confidence intervals, and shaded areas indicate 1***σ*** standard deviations. Similar SHLs between SjD+ and control groups, diverging from NHANES, suggest testing bias.

Serum hormone levels (SHLs) were consistent between SjD and control groups. **Figure 1C** shows serum hormone levels (SHLs) for testosterone, estradiol, and SHBG in the SjD, control, and NHANES cohorts. A slight reduction in testosterone was noted in SjD males aged 40–60 compared to controls, though this effect was marginal (Cohen’s d ≈ 0.5) and not significant. SHBG and estradiol levels also showed minimal variation from controls, with overlapping confidence intervals across age groups. Differences were observed between both the male and female cohorts and the NHANES cohort, suggesting evidence of a testing bias. These results suggest that deviations in testosterone, estradiol, or SHBG are unlikely to be primary factors in SjD development. Instead, the observed prevalence patterns more likely reflect the influence of normal hormone fluctuations, age-related immune modulation, and other mechanisms underlying disease pathogenesis.

Best-fit hormonal models for the SPF confirmed a strong correlation between SPF and SHLs of both estradiol and testosterone. Goodness-of-fit metrics were comparable between the two hormones, with estradiol showing a marginally better fit (AIC: 21.43 for estradiol vs. 21.69 for testosterone; BIC: -111.37 for estradiol vs. - 111.11 for testosterone) (**Supplemental Figure 1**). Additionally, the residuals were normally distributed for both models, indicating that they were unbiased and accurately captured the trends in the data. These results further support the hypothesis that typical levels of sex hormones contribute to the modulation of sex-specific SjD prevalence across different age groups.

## Discussion

Our findings highlight a strong association between age-related hormonal modulation and sex-specific SjD prevalence. Rather than viewing the sex bias in SjD as static, our data suggest an evolving interplay between testosterone, estradiol, and SHBG levels across different life stages, modulating disease susceptibility. This perspective challenges traditional models that consider sex bias as a fixed prevalence and instead frames it as age-dependent, influenced by natural hormonal fluctuations rather than deviations from expected levels.

Our study identifies a striking shift in the prevalence of SjD among males, with pediatric SjD patients exhibiting a male prevalence of 30.06%, which sharply declines to 9.85% in adults. This dramatic shift highlights the potential role of age-related hormonal changes, particularly androgens, in modulating disease susceptibility. Prior studies have noted a higher male prevalence in pediatric SjD, but with smaller sample sizes and a less pronounced sex disparity. For instance, Ramos-Casals et al. (2021) characterized a pediatric SjD cohort of 158 children, reporting 13.9% male prevalence^12^, while Basiaga et al. (2021) analyzed 300 pediatric SjD cases, identifying 17% male prevalence^13^. Additionally, Means et al. (2017) systematically reviewed pediatric SjD cases, with individual studies reporting male prevalence ranging from 15–25%^14^. Our findings, based on a significantly larger cohort, provide stronger epidemiological evidence for this sex-based shift and further support the hypothesis that hormonal regulation may play a key role in SjD pathogenesis.

There is compelling evidence that testosterone and other androgens may play a protective role in SjD. During puberty, when testosterone levels surge^15^, we observed a marked decrease in SjD prevalence in males, suggesting a dose-dependent protective effect of androgens. Conversely, as testosterone levels decline with age, there is a gradual increase in SjD prevalence, reinforcing the role of androgens in immune modulation^16^. Further supporting this hypothesis, males with Klinefelter syndrome (47,XXY), who have inherently lower androgen levels, exhibit autoimmune profiles more similar to females, including increased susceptibility to SjD^17,18^. Notably, testosterone therapy in these individuals has been associated with remission of SjD symptoms. While past androgen therapy trials in SjD have yielded mixed results^19–22^, this may be due to challenges in achieving the correct balance of testosterone, estrogen, and free SHBG and ability to target delivery to disease affected tissues rather than a lack of efficacy. Additionally, other factors such as chronic inflammation, epigenetic changes, and autoantibody-mediated tissue damage may contribute to immune dysregulation beyond hormonal effects.

The testosterone-to-estrogen ratio appears to be a critical factor in SjD susceptibility, particularly in women during menopause, when hormone levels undergo a drastic shift. In adult women, testosterone declines steadily, while estrogen levels remain relatively stable until menopause, when they sharply decrease^23^. This sudden estrogen drop alters the testosterone-to-estrogen ratio, increasing relative androgenic influence. However, this shift may not be beneficial if free testosterone is concurrently reduced by increasing SHBG levels, as testosterone binds to SHBG with higher affinity compared to estradiol^24^. SHBG follows a U-shaped concentration curve across the lifespan, declining from early adulthood to midlife but rising again in later age^25^. Because estrogen stimulates SHBG production^26^, the sharp decline in estrogen during menopause initially reduces SHBG levels. However, as aging progresses, SHBG levels increase^25^, further binding free testosterone and reducing its bioavailability. If testosterone is protective against SjD, then this combined effect of increasing SHBG and declining estrogen may lead to a functional decrease in available protective testosterone during menopause, contributing to the higher prevalence of SjD in postmenopausal women. These findings suggest that modulating testosterone-to-estrogen ratios or addressing SHBG-mediated reductions in free androgens could represent a novel therapeutic approach for SjD prevention and management.

While robust, our study has limitations. SjD diagnosis criteria may differ across healthcare providers, as this study relies on provider-based diagnoses. Hormone profiles are often only measured in individuals suspected of endocrine abnormalities, potentially introducing selection bias. Additionally, systemic hormone levels may not fully capture tissue-specific hormonal activity, particularly within exocrine glands relevant to SjD pathogenesis. Survivorship bias may also play a role, as individuals with longer disease durations or more frequent healthcare visits may have higher rates of hormone testing. Despite these limitations, our findings provide strong evidence for hormone-mediated susceptibility in SjD, underscoring the importance of further research into hormonal influences on autoimmunity.

Future research should focus on longitudinal studies tracking hormone fluctuations over time to clarify their role in immune modulation and SjD pathogenesis^11^. Additionally, assessing SHBG and bioavailable testosterone levels in SjD patients at different life stages may offer further insight into the mechanistic underpinnings of sex hormone influence on autoimmunity. Clinically, our findings suggest that considering hormone levels in personalized SjD treatment strategies may improve alignment with natural hormonal shifts that contribute to disease susceptibility.

## Data Availability

All data produced are available through request to TriNetX, LLC.

https://trinetx.com/

## Acknowledgements

We acknowledge the support and resources provided by TriNetX for EHR access.

