## Supplemental Material for "Age-Dependent Sex Disparities in Sjogren’s Disease Prevalence Align with Natural Hormone Fluctuations"

### Methods

This study focuses on cohort design, data processing, and statistical modeling to analyze the relationship between hormone profiles and SjD prevalence. The study was conducted in accordance with the ethical standards set by the Institutional Review Board (IRB), managed by the Office of Research Integrity and Compliance.

#### Cohort Construction and Data Preparation

Patient electronic health records (EHRs) were obtained on September 19th, 2023, from the TriNetX Research Network (TNXRN), which provides access to electronic medical records containing diagnoses, procedures, medications, laboratory values, and genomic data from approximately 120 million de-identified records across 83 healthcare organizations, using select ICD-10-CM: M35.03, Sjogren's syndrome with myopathy;  M35.04, Sjogren's syndrome with tubulointerstitial nephropathy; M35.02, Sjogren's syndrome with lung involvement; M35.01, Sjogren's syndrome with keratoconjunctivitis; M35.00, Sjogren's syndrome, unspecified; M35.0, Sjogren's syndrome; M35.06, Sjogren's syndrome with nervous system involvement; M35.05, Sjogren's syndrome with inflammatory arthritis; M35.09, Sjogren's syndrome with other organ involvement; M35.0C , Sjogren's syndrome with dental involvement; M35.0B, Sjogren's syndrome with vasculitis; M35.0A, Sjogren's syndrome with glomerular disease; M35.07, Sjogren's syndrome with central nervous system involvement; M35.08, Sjogren's syndrome with gastrointestinal involvement; H16.2, Keratoconjunctivitis; H16.22, Keratoconjunctivitis sicca, not specified as Sjogren's; H16.229, Keratoconjunctivitis sicca, not specified as Sjogren's, unspecified eye; H16.223, Keratoconjunctivitis sicca, not specified as SjD, bilateral; H16.20, Unspecified Keratoconjunctivitis; H04.12, Dry Eye Syndrome; R86.2, Dry Mouth, unspecified.

The SjD cohort (N=101,856) included patients with at least two M35.0 ICD-10-CM codes (or subcodes) spaced by at least six months to ensure diagnostic accuracy. Control groups included patients with SjD-associated symptoms but no diagnosis (dry eye, dry mouth, or keratoconjunctivitis; N=1,333,082 total). Each patient's diagnosis date (t) was defined as the earliest relevant ICD-10-CM code entry. Serum hormone levels (testosterone, estradiol, and sex hormone-binding globulin [SHBG]) were extracted using LOINC codes with the test closest to the diagnosis date used for analysis. Although testing rates were low, the dataset size ensured sufficient statistical power. Testosterone (CPT 2986-8, Testosterone [Mass/volume] in Serum or Plasma; CPT 14913-8, Testosterone [Moles/volume] in Serum or Plasma; CPT 83116-4, Testosterone [Mass/volume] in Serum or Plasma by Immunoassay; CPT 105121-8,Testosterone [Mass/volume] in Serum or Plasma by LC/MS/MS; CPT 83115-6, Testosterone [Moles/volume] in Serum or Plasma by Immunoassay; CPT 49041-7, Testosterone [Mass/volume] in Serum or Plasma by Detection limit <= 1.0 ng/dL; CPT 70239-9, Testosterone [Moles/volume] in Serum or Plasma by Detection limit <= 3.47 pmol/L), Estrogen (CPT 2254-1, Estrogen [Mass/volume] in Serum or Plasma; CPT 70207-6, Estrogen [Moles/volume] in Serum or Plasm; CPT 53765-4, Estrogen [Mass/volume] in Serum or Plasma by calculation, CPT 2243-4, Estradiol (E2) [Mass/volume] in Serum or Plasma; CPT 14715-7, Estradiol (E2) [Moles/volume] in Serum or Plasma; CPT 72873-3, Estradiol (E2) [Moles/volume] in Serum or Plasma by High sensitivity method; CPT 35384-7, Estradiol (E2) [Mass/volume] in Serum or Plasma by High sensitivity method; CPT 83096-8, Estradiol (E2) [Mass/volume] in Serum or Plasma by Immunoassay; CPT 83097-6, Estradiol (E2) [Moles/volume] in Serum or Plasma by Immunoassay), SHBG (CPT 2942-1, Sex hormone binding globulin [Mass/volume] in Serum or Plasma; CPT 13967-5, Sex hormone binding globulin [Moles/volume] in Serum or Plasma).

Each patient's diagnosis date (t) was defined as the earliest relevant ICD-10-CM code entry. Serum hormone levels (testosterone, estradiol, and sex hormone-binding globulin (SHBG)) was extracted using LOINC codes, with the test closest to the diagnosis date used for analysis. In the SjD group, testing rates were 4.10% for testosterone, 4.06% for estradiol, and 1.15% for SHBG, while the control group had rates of 4.81%, 2.34%, and 1.64%, respectively. Despite low testing rates, the scale of the data provided sufficient statistical power.

### Population Baseline and Imputation Using NHANES Data

Routine hormone testing was infrequent in both the SjD and control cohorts, leading to a bias toward abnormal serum hormone levels (SHLs). Since testing standards were consistent across SjD+ and Control (C) groups, this bias was assumed to be the same. To correct this, NHANES datasets (2013–14, 2015–16 cycles) were used to provide population-level baseline SHLs.

Patients in each dataset (SjD, C, and NHANES), patients were divided by sex and binned in 5-year increments based on patient age at diagnosis / testing (PAaD). SHLs in each bin were modeled as log-normal distributions, with the mean (μ) and standard deviation (σ) estimated using maximum likelihood estimation (MLE). Welch's *t*-test was applied to compare SHL means between cohorts under the null hypothesis of equal means. SHLs in the SjD and control groups showed minimal statistically significant variation, confirming that SjD patients do not exhibit hormone disruptions at the population level. In regions where SjD and control were dissimilar, the effect size was calculated using Cohen-*d* [27], which indicated maximum effect sizes on the order of 0.5. This aligns with existing literature [7]. However, significant differences between SjD and NHANES suggest a testing bias as all subjects in the NHANES data undergo testing independent of suspected disease state.

To address the testing bias in the SjD group, we imputed missing hormone data for each patient by randomly selecting hormone values from the corresponding NHANES age and sex bin. This method ensured that each patient was assigned a representative hormone value, minimizing bias due to incomplete data while preserving any abnormal values directly reported in the EHRs. By leveraging population-level data, this imputation approach effectively corrected for bias, providing a more accurate reflection of baseline hormone levels across the cohort. Notably, this approach relies explicitly on the homogeneity between the SjD and control groups.

### Male Sex Prevalence

To quantify the sex bias in the SjD population, we computed the male sex prevalence fraction (SPF), representing the proportion of male patients diagnosed at each age. Patients were grouped into bins based on patient age at diagnosis (PAaD), with a bin width of 2.5 years. The SPF for each bin was modeled as a binomially distributed random variable, reflecting the probability of a patient being male within that age group. The SPF was estimated using the maximum a posteriori (MAP) estimator with uninformative priors to avoid biasing the results.

Figure 1 presents the calculated SPF and its 95% credible intervals, providing a robust measure of uncertainty. This approach enables a reliable assessment of male prevalence across age groups, characterizing the sex bias in SjD+ patients.

### Analysis of Hormone Profiles and Sex Ratios

To investigate the relationship between SHLs and the SPF, we employed a generalized linear model (GLM), where the SPF was modeled as a function of age at diagnosis (τ) and individual (male) hormone levels ($H$), denoted as $Y \sim \log H$. This approach was chosen to assess how variations in hormone levels might influence the observed sex distribution in SjD, based on the hypothesis that hormone profiles contribute to the gender-specific patterns in disease prevalence.

A logit link function accounted for the binary nature of the sex data, and GLMs were fit using iteratively reweighted least squares (IRLS) to handle the non-linear relationships inherent in a binomial framework. Model performance was evaluated using the Akaike Information Criterion (AIC) and Bayesian Information Criterion (BIC), allowing for comparison across hormone-specific models.

**Supplemental Figure 1. Comparison of SPF and SHLs in SjD+ patients by age at diagnosis.** Each row presents data for testosterone, estradiol (E2), and SHBG, respectively. **(A)** Male SPF in SjD+ patients (black) as a function of age at diagnosis, compared with the hormone SHL function (blue). The SPF curve shows the maximum a posteriori (MAP) estimate per age bin, with 95% credible intervals as error bars. Mean SHLs are estimated by maximum likelihood (MLE) under a log-normal distribution, with error bars indicating 3 standard errors of the mean (SEM). **(B)** Each hormone's SPF curve is compared to its best-fit generalized linear model (GLM), with shaded areas representing the standard error of the model's prediction. **(C)** Residuals and their distributions for each hormone's model, with well-fitting models expected to yield residuals normally distributed around zero. Comparative quality statistics (BIC and AIC) are provided. Patients are binned in 2.5-year intervals from ages 5 to 90.

Supplemental Figure 1.

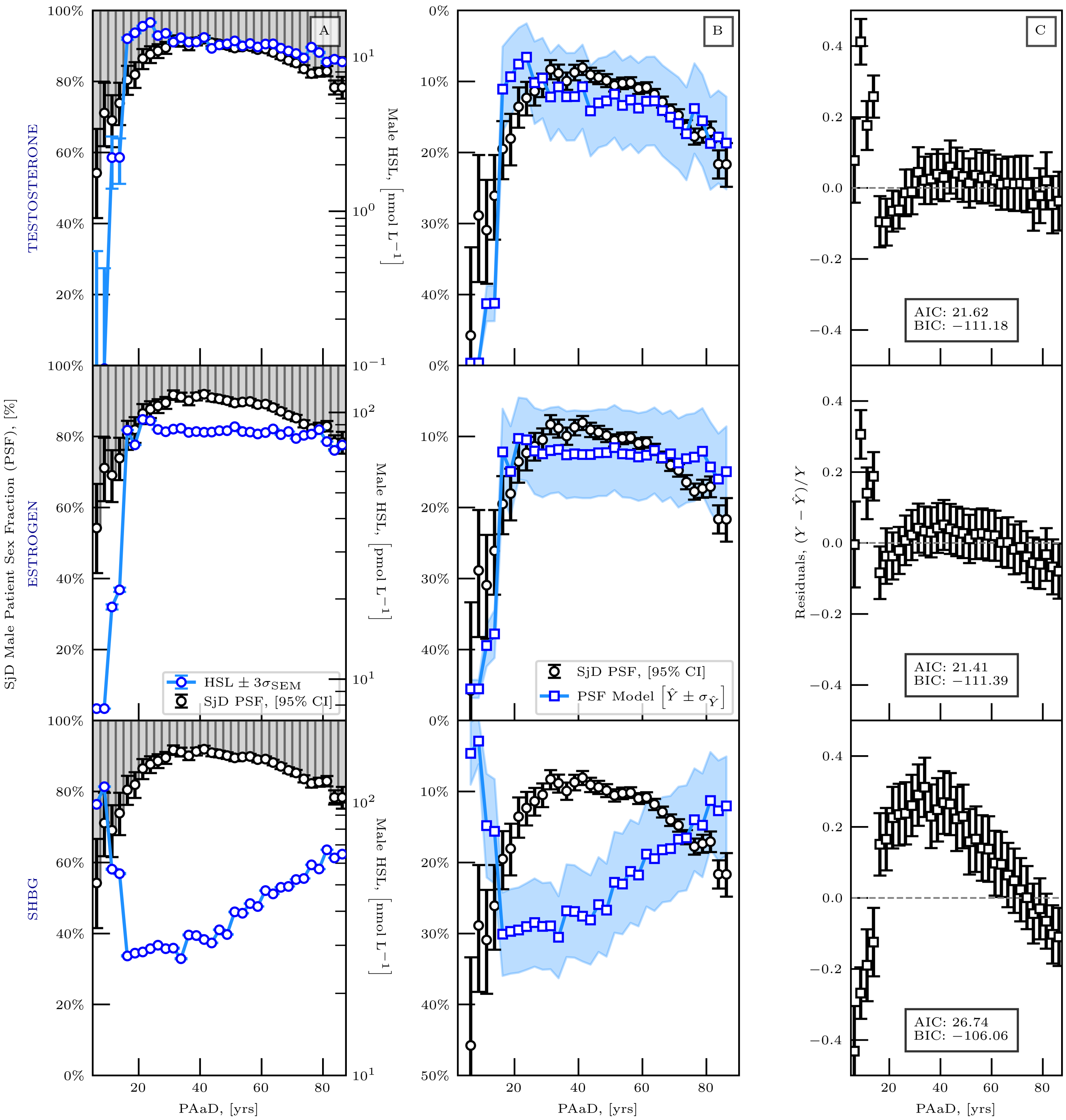
